# Identification of abnormal patterns of evolution of neurocysticercosis cysts and what they might indicate

**DOI:** 10.1101/2025.07.12.25331432

**Authors:** Meghana G. Shamsunder, Afroza Parvin, Hongbin Zhang, Arturo Carpio, W. Allen Hauser, Alex Jaramillo, Jorge Moncayo, Karina Quinde Herrera, Elizabeth A. Kelvin

**Author notes:** Corresponding author: Elizabeth A. Kelvin, 55 West 125^th^ Street New York, NY 10027.

## Abstract

Few studies have described unexpected patterns of cyst evolution for neurocysticercosis (NCC), an infection of the central nervous system with the larval stage of the pork tapeworm, *Taenia solium*. We used data from a randomized controlled trial on albendazole treatment, conducted in Ecuador and including 178 NCC patients who had imaging at months one, six, 12, and 24 to examine unexpected patterns of NCC cyst evolution by potential explanation. Over half (57.2%) of patients exhibited an unusual pattern of cyst evolution. The most common pattern was consistent with possible cyst migration (44.8%), followed by reverse transitioning of NCC cysts to earlier phases (16.6%). Only 1.4% had evidence suggesting a new infection. Documenting unexpected patterns of NCC cyst evolution and exploring potential explanations for these patterns may inform radiologists’ reading of NCC patient scans to improve care and generate future research ideas to help us better understand this parasite and disease.

**Article Summary Line:** We documented several different unexpected patterns of NCC cyst evolution and explored potential explanations for these patterns. This may inform radiologists’ reading of NCC patient scans to improve care.

## Introduction

Neurocysticercosis (NCC) is a parasitic infection of the central nervous system (CNS) with the larval stage of the pork tapeworm, *Taenia solium.* NCC is a leading cause of acute symptomatic seizures, particularly in low- and middle-income countries, however, NCC diagnosis is increasing in the United States.[1–6] Despite this, NCC is classified as a neglected disease by both the Centers for Disease Control[7] and the World Health Organization,[8] and the life course of encysted NCC larva in the CNS remains poorly understood. Research suggests that these cysts evolve in the human brain through three distinct phases: (1) an active phase, where the parasite is viable/alive and is capable of hiding from the host immune system by surrounding itself with immunomodulatory host immunoglobulins, often resulting in minimal symptoms in the host; (2) a transitional phase, in which the parasite is degenerating and has been recognized by the host immune system, resulting in inflammation around the cyst often causing more severe symptoms; and, although some NCC cysts completely resolve and are no longer detectable on imaging, others leave a (3) residual calcification.[9–11] In some cases, calcified cysts may continue to elicit a host immune response,[12] and can be associated with continued symptoms.[13]

There are many challenges to understanding NCC cyst evolution. For one, it remains unclear how long cysts remain in each phase and what factors influence evolution.[14] A number of studies, including randomized controlled trials (RCT), indicate that treatment with albendazole hastens larval death and cyst evolution from the active to transitional phase,[4,15] making albendazole a standard treatment for this infection.[3,4,13,16] However, albendazole treatment alone results in parasitic death for approximately 40% of patients,[4] with evidence of differential treatment effects depending on the cyst location and the presence and phase of other cysts within the brain.[5,16] Host characteristics such as age, gender, and immune system functioning also appear to play a role in NCC cyst evolution.[17–20]

Imaging type further complicates our understanding of cyst evolution. Computed tomography (CT) better distinguish calcified cysts while magnetic resonance imaging (MRI) is suited for identifying active and transitional cysts.[10,11,21,22] Thus, patient variability in cyst evolution may be due to differences in the form of imaging used to follow their infection. Compounding this, the inter-rater reliability of radiological readings generally tends to be mediocre.[23–25] In one study, the kappas comparing readings from two radiologists identifying NCC cysts in specific phases and brain locations (parenchymal versus extraparencymal) were only fair to good (0.4-0.7).[4] Related to imaging, the presence of localized inflammation and edema may lead to challenges in identification since “new” NCC cysts may become visible on imaging once inflammation resolves (e.g. after administering corticosteroids). Edema is very common for transitional cysts[26] and has also been described for calcified cysts.[27–29]

Other complications include potential reinfection among those with NCC cysts. New infections may either lead to an increase in cyst burden (documented on imaging) or may be misclassified as a previous cyst that resolved. There has also been some suggestion that NCC cysts have the capacity of gemmation (asexual reproduction by budding), especially in the subarachnoid space,[26] and that cysts may migrate to other locations in the brain, resulting in the appearance of cyst resolution in one location but a new infection in a different location.[30] Lastly, although it is assumed that NCC cysts can only evolve in one direction, one study following individual cysts over time found 14 cysts that appeared to reverse transition.[5] This was thought to be due to human error in reading images, yet it remains theoretically possible that a transitional cyst might fight the host immune system and return to the active phase.

NCC cysts are usually followed in aggregate, where cysts in each phase within the overall brain and in specific locations are counted and tracked as a group. This makes it easy to miss unexpected patterns of cyst evolution. Given the numerous complications to understanding this neglected infectious disease, description of cyst evolutionary patterns is a key to moving research on this infection forward. Understanding cyst evolution is important to both guiding the reading of NCC imaging accurately and understanding the benefits and shortcomings of current treatments. Therefore, the aims of this study were to describe unexpected patterns of NCC cyst evolution based on radiological readings of patients enrolled in a RCT of albendazole treatment followed over two years. We categorized the unexpected patterns over time by possible explanations that complicate our understanding of NCC. We hope that our description of these patterns will improve current understanding about possible evolutionary paths of *T. solium* larva in human brains.

## Methods

### Data Source and Study Design

Data came from an RCT conducted in 2001-2003 (registered at ClinicalTrials.gov, # NCT00283699) that assessed the impact of albendazole treatment versus placebo on NCC cyst resolution among 178 patients diagnosed with active or transitional NCC cysts recruited from six hospitals in Ecuador. The study has been previously described[4] and was noted as one of only two high quality trials on treatment for NCC.[16] Here we provide a brief description. Patients were randomized to treatment with albendazole plus prednisone or placebo plus prednisone. Albendazole or placebo was given orally every 12 hours for eight days under direct observation. Prednisone was taken for three weeks at decreasing doses. CT or MRI scans were conducted at baseline and follow-up months one, six, 12, and 24. For logistical reasons, most scans at month 24 were CT and many patients transitioned from MRI to CT at this time point. Data was collected in aggregate to describe the total number of cysts in the active, transitional, and calcified phase in the brain overall and in each brain-location.

### Sample for Analysis

For these analyses we included a subset of the 178 participants, specifically those who had a single cyst in one or more brain locations at baseline. Although these participants may have multiple NCC cysts in other brain locations, we described evolutionary patterns for those brain locations with a single cyst. Among this subset of patients with at least one brain location having a single NCC cyst, we characterized the total burden of cysts in the entire brain when describing the abnormal patterns. The total number of participants who met the criteria for inclusion (i.e., having at least one brain location with a single NCC cyst at baseline) was 145, for whom we followed 378 individual cysts through 1,372 scans through baseline, months one, six, 12 and/or 24 (note 518 scans were missing at one or more follow-up times). To note, while the baseline scan had only one cyst, later scans may have had more than one cyst.

### Identification of Abnormal Patterns of Cyst Evolution and Possible Explanations

Based on previous research, NCC cyst evolution is expected to follow these patterns:

1. Evolution is expected to be unidirectional (no reverse transitions); the total number of cysts in each phase within a patient’s brain is expected to, (1) remain the same in the next scan, (2) resolve in earlier phases (decrease in cysts in earlier phases), (3) evolve from earlier phases to later phases resulting in an increase in cysts in later phases. For example, if there was an increase in the number of calcified cysts from two to four between months one and six, we would expect to see a reduction in the number of active and/or transitional cysts by at least two during the same period (the decrease could be greater than two if some cysts completely resolved).
2. The number of cysts in the brain, overall, and within a specific brain location will either stay the same or decrease over time (i.e., we do not expect overall cyst burden nor burden in a specific location to increase over time). While it remains unclear to what extent NCC patients are at risk for reinfection,[31] one study measuring reinfection by serology testing suggests that reinfection is rare (0.5-3.7%).[32]

### Measures

Based on the expectations above, we looked for unexpected patterns of cyst evolution and evidence consistent with possible explanations for these patterns (Table 1). To note, all unexpected patterns can be due to human error in reading the scans. The unexpected patterns we looked for included:

1. Reverse transitioning defined as across scans in locations with a baseline total of one cyst, there was a decrease in cysts in later phases and simultaneous increase in cysts in earlier phases (suggesting a reverse transition) with no change in the total number of cysts in the patient’s brain (suggesting that the increase in cysts in an earlier phase was not a new infection). Other studies have considered reverse transitioning of NCC cysts but it has neither been well documented nor explained.[5,33]
2. Compared to the previous scan, an increase in the total number of cysts in a specific brain location with an increase or no change in cyst burden in the overall brain. The explanation for this pattern could include:

a. Cyst migration defined as across scans in locations with a baseline total of one cyst, there was an increase in cysts in that location and no change in the total number of cysts in patient’s brain. A cyst may have “moved” from an adjacent location, leading to an apparent increase in one region with a reduction in another region, a pattern which has been described previously.[2,30] For example, if the total number of cysts in the right temporal lobe increased <u>and</u> the total number of cysts in the right frontal lobe decreased all while the total brain burden stayed the same, this might be evidence of cyst migration.
b. New infection defined as across scans in locations with a baseline total of one cyst, there was an increase in active cysts in that location and an increase in the total number of cysts in the brain. For example, if a patient had one active and one transitional cyst in their brain, then in the next scan s/he had two active and one transitional cyst this increase might be due to a new infection.
c. Change in imaging device defined as across scans in locations with a baseline total of one cyst, an increase in calcified cysts due to changing from MRI to CT. For example, if a patient had two active and two transitional cysts evident on MRI but three transitional and two calcified cysts in the next visit <u>and</u> imaging changed to a CT, it is possible that a calcified cyst was not detected earlier by the MRI but became apparent in the CT scan (since CT scans are better for detecting calcified cysts).[20,26,31]
d. Abnormal patterns not easily explained were defined as an increase in cysts within locations with a baseline total of one cyst <u>and</u> where the patient did not experience a change in imaging device.

**Table 1:** Description of abnormal patterns of neurocysticercosis cyst evolution.

| Explanation | Pattern |  | Alternate explanations |
| --- | --- | --- | --- |
|  | In Specific Brain Location with a Single Cyst at Baseline | In the Overall Brain |  |
| **Reverse Transition** | Three reverse transitions were assessed:<br>1. Calcified to Transitional<br>2. Transitional to Active<br>3. Calcified to Active<br>For each of these, we looked at a decrease in the number of cysts in the later phase with a simultaneous increase in cysts in the earlier phase | No change in the total number of cysts in the brain | 1. The original cyst resolved and was replaced with a new infection (i.e., the cyst in an earlier phase in the same location)<br>2. Error in radiologist reading of one of the scans |
| **Migration** | Increase in the number of cysts in a specific location with one cyst at baseline to >1 | No change in the total number of cysts in the brain | 1. Some cysts in the brain may have resolved and the new cyst is a new infection<br>2. Error in radiologist reading of one of the scans |
| **New infection** | An increase in the number of active cysts in the location being followed | An increase in the total number of cysts in the brain. | 1. Error in radiologist reading of one of the scans |
| **Increase in calcified cysts with change from MRI to CT scan at 24 months** | Increase in the number of calcified cysts in a location between 12 and 24 months among patients who had an MRI at month 12 and a CT scan at month 24 | Either no change in the total number of cysts in the brain (some cysts may have resolved) or an increase in the total number of cysts in the brain between 12 and 24 months | 1. Error in radiologist reading of one of the scans |
| **Abnormal patterns that cannot be easily explained: Increase in cyst of any phase with no change in image device** | An increase in the number of active, transitional, or calcified cysts in the location being followed where image device was not changed | Either no change in the total number of cysts in the brain (some cysts may have resolved) or an increase in the total number of cysts in the brain | 1. Brain swelling and edema subsiding leading to existing cysts becoming visible<br>2. Error in radiologist reading of one of the scans |

We also described participants in terms of treatment (albendazole versus placebo), and demographic characteristics (patient age in years, sex, and race/ethnicity according to the categories used in Ecuador).

### Statistical analysis

We describe the study participants overall and by presence/absence of at least one of the abnormal cyst patterns described above. We assessed the statistical significance with either a Mann-Whitney (for age) or Fishers Exact Test (for categorical variables). We described the number of cases of each of the abnormal patterns described above. All analyses were conducted in R Statistical Software (Vienna, Austria) and statistical significance was set at α=0.05.

## Results

### Description of the Participants & Cyst Followed

Of the 178 patients included in the RCT, 145 participants were included in these analyses who had scans with one NCC cyst in one or more specific brain locations. Overall, 67 (46.2%) participants were treated with albendazole and 78 (53.8%) with placebo. The average age of participants was 40.7 years, 55.9% were male, and the majority identified as Mestizo (88.3%). There were no statistical differences by presence of one or more abnormal cyst patterns. (Table 2).

**Table 2:** Demographics and Cyst Related Characteristics at the Patient Level.

|  | Total | Patients with no abnormal patterns of cyst evolution | Patients with one or more abnormal pattern of cyst evolution | P-value |
| --- | --- | --- | --- | --- |
| **Demographics** |  |  |  |  |
| **Total, n (row %)** | 145 | 62 (42.8) | 83 (57.2) | NA |
| **Treatment, n (column %)\*** |  |  |  | 0.4 |
| Albendazole | 67 (46.2) | 26 (41.9) | 41 (49.4) |  |
| Control | 78 (53.8) | 36 (58.1) | 42 (50.6) |  |
| **Age (years)** |  |  |  | 0.9 |
| Mean (SD) | 40.7 (17.4) | 40.9 (18.8) | 40.5 (16.3) |  |
| Median (IQR) | 40 (28.6, 50.8) | 38.5 (25.2, 54.6) | 40.3 (28.9, 49.5) |  |
| **Sex, n (column %)\*** |  |  |  | 0.4 |
| Male | 81 (55.9) | 32 (51.6) | 49 (59) |  |
| Female | 64 (44.1) | 30 (48.4) | 34 (41) |  |
| **Race, n (column %)\*** |  |  |  | 0.3 |
| Mestizo | 128 (88.3) | 53 (85.5) | 75 (90.4) |  |
| Indigenous | 1 (0.7) | 1 (1.6) | 0 (0) |  |
| Black | 2 (1.4) | 2 (3.2) | 0 (0) |  |
| White | 14 (9.7) | 6 (9.7) | 8 (9.6) |  |
| **Cyst abnormalities by potential explanation** |  |  |  |  |
| **Reverse transitioning, n (column %)\*** | 24 (16.6) |  | 24 (28.9) |  |
| **Cyst migration, n (column %)\*** | 65 (44.8) |  | 65 (78.3) |  |
| **New infection, n (column %)\*** | 2 (1.4) |  | 2 (2.4) |  |
| **Increase in calcified cysts with change from MRI to CT scan at 24 months, n (column %)\*** | 12 (8.3) |  | 12 (14.5) |  |
| **Abnormal patterns that cannot be easily explained, n (column %)\*** | 60 (41.4) |  | 60 (72.3) |  |
| Fisher's Exact Test used for categorical variables, Mann-Whitney U test used for age comparisons |  |  |  |  |
| * Denominator is total number of patients by presence/absence of abnormal pattern (i.e., column percents). |  |  |  |  |

These 145 patients had a total of 378 cysts that were the only cyst in a particular brain location at baseline, and these cysts were followed through 1372 scans. These cysts were in 11 different brain locations. (Table 3) Among the 145 patients, 83 (57.2%) individuals had evidence of one or more abnormal patterns of cyst evolution with 24 (16.6%) patients having evidence of possible reverse transitions, 65 (44.8%) showing evidence of a possible cyst migration, 2 (1.4%) with evidence of a possible new infection, 12 (8.3%) with new calcified cysts potentially due to changing from an MRI at 12 months to a CT scan at 24 months, and 60 (41.4%) with abnormal patterns that cannot be easily explained. (Table 2) Across the 1372 scans, there was evidence suggesting reverse transition in 31 (2.3%) scans, cyst migration in 141 (10.3%) of scans, new infection in 2 (0.1%) scans, calcified cysts appearing due to a change from MRI to CT scan in 14 (1.0%) scans, and 113 (29.9%) scans with abnormal patterns not easily explained. (Table 3)

**Table 3:** Description of Cysts Followed Through Imaging and Scan Information.

|  | Total | Patients with no abnormal patterns of cyst evolution | Patients with one or more abnormal pattern of cyst evolution |
| --- | --- | --- | --- |
| **Number of cysts in locations with only 1 cyst at baseline, n (row %)** | 378 | 230 (60.8) | 148 (39.2) |
| **Location of cysts followed, n (column %)<sup>\*</sup>** |  |  |  |
| Frontal Lobe | 86 (22.8) | 57 (24.8) | 29 (19.6) |
| Parietal lobe | 79 (20.9) | 42 (18.3) | 37 (25) |
| Temporal lobe | 68 (18.0) | 40 (17.4) | 28 (18.9) |
| Occipital lobe | 44 (11.6) | 27 (11.7) | 17 (11.5) |
| Basal ganglia & Internal capsule | 37 (9.8) | 21 (9.1) | 16 (10.8) |
| Posterior fossa | 19 (5.0) | 12 (5.2) | 7 (4.7) |
| Sylvian Fissure | 18 (4.8) | 13 (5.7) | 5 (3.4) |
| Other Cisternal Locations | 13 (3.4) | 9 (3.9) | 4 (2.7) |
| IV ventricle | 7 (1.9) | 5 (2.2) | 2 (1.4) |
| Lateral Ventricles | 6 (1.6) | 3 (1.3) | 3 (2) |
| III Ventricle | 1 (0.3) | 1 (0.4) | 0 (0) |
| **Total Number of Scans, n (row %)** | 1372 | 719 (52.4) | 653 (47.6) |
| **Scans available at each follow-up, n (column %)<sup>†</sup>** |  |  |  |
| Baseline | 378 (27.6) | 230 (100) | 148 (100) |
| Month 1 | 377 (27.5) | 229 (99.6) | 148 (100) |
| Month 6 | 263 (19.2) | 130 (56.5) | 133 (89.9) |
| Month 12 | 226 (16.5) | 96 (41.7) | 130 (87.8) |
| Month 24 | 128 (9.3) | 34 (14.8) | 94 (63.5) |
| **Abnormal patterns observed in scans, n (column %)<sup>†</sup>** |  |  |  |
| Reverse Transition | 31 (2.3) |  | 31 (20.9) |
| Migration | 141 (10.3) |  | 141 (95.3) |
| New Infection | 2 (0.1) |  | 2 (1.4) |
| Increase in calcified cysts with change from MRI to CT scan at 24 months | 14 (1.0) |  | 14 (9.5) |
| Abnormal patterns that cannot be easily explained | 113 (29.9) |  | 113 (76.4) |

\* Denominator is number of cysts in location with only 1 cyst at baseline by patient presence/absence of abnormal pattern. † Denominator is number of scans by patients with/without abnormal patterns.

### Abnormal Patterns in Detail (Table 4)

Among the 31 scans (2.3%) with evidence consistent with reverse transitions, 19 (61.3%) indicated a transition from a transitional to active cyst, 7 (22.6%) from calcified to transitional cyst, and 5 (16.1%) from calcified to active cyst. Most of these reverse transitions occurred in the frontal (35.5%), temporal (19.4%) and occipital (12.9%) lobes, with 22.6% occurring at month one, 32.3% at month six, 16.1% at month 12, and 29.0% at month 24 of follow-up.

**Table 4:** Abnormal Patterns Observed for the 378 cysts followed in 1372 Scans in 145 Patients.

| Pattern and Description |  | Change in cysts in the brain, overall | Number of scans with this pattern, n (%) | Lobes and Locations Where Pattern was Identified, n (row %)<sup>†</sup> |  | Follow-up Time when Pattern was Identified, n (row %) |  |
| --- | --- | --- | --- | --- | --- | --- | --- |
| 1. | Reverse Transition | No Change | 31 (2.3)<sup>†</sup> | Frontal<br>Temporal<br>Occipital<br>Parietal<br>Sylvian Fissure Cistern<br>Other Cisternal Location<br>Basal Ganglia & Internal Capsule<br>IV Ventricle<br>Posterior Fossa | 11 (35.5)<br>6 (19.4)<br>4 (12.9)<br>3 (9.7)<br>2 (6.5)<br>2 (6.5)<br>1 (3.2)<br>1 (3.2)<br>1 (3.2) | Month 1<br>Month 6<br>Month 12<br>Month 24 | 7 (22.6)<br>10 (32.3)<br>5 (16.1)<br>9 (29.0) |
| 1a. | ↑ Transitional &<br>↓ Calcified cysts | No Change | 7 (22.6)<sup>‡</sup> | Temporal<br>Frontal<br>Occipital<br>Basal Ganglia & Internal Capsule | 3 (42.9)<br>2 (28.6)<br>1 (14.3)<br>1 (14.3) | Month 1<br>Month 6<br>Month 12<br>Month 24 | 2 (28.6)<br>1 (14.3)<br>1 (14.3)<br>3 (42.9) |
| 1b. | ↑ Active &<br>↓ Transitional cysts | No Change | 19 (61.3)<sup>‡</sup> | Frontal<br>Parietal<br>Occipital<br>Temporal<br>Sylvian Fissure Cistern<br>IV Ventricle<br>Posterior Fossa<br>Other Cisternal Location | 6 (31.6)<br>3 (15.8)<br>3 (15.8)<br>2 (10.5)<br>2 (10.5)<br>1 (5.3)<br>1 (5.3)<br>1 (5.3) | Month 1<br>Month 6<br>Month 12<br>Month 24 | 5 (26.3)<br>7 (36.8)<br>4 (21.1)<br>3 (15.8) |
| 1c. | ↑ Active &<br>↓ Calcified cysts | No Change | 5 (16.1)<sup>‡</sup> | Frontal<br>Temporal<br>Other Cisternal Location | 3 (60.0)<br>1 (20.0)<br>1 (20.0) | Month 6<br>Month 24 | 2 (40.0)<br>3 (60.0) |
| 2. | Migration (↑ in Location) | No Change | 141 (10.3)<sup>†</sup> | Parietal<br>Temporal<br>Frontal<br>Basal Ganglia & Internal Capsule<br>Occipital<br>Posterior Fossa<br>Lateral Ventricles<br>Other Cisternal Location<br>IV Ventricle<br>Sylvian Fissure Cistern | 42 (29.8)<br>33 (23.4)<br>23 (16.3)<br>16 (11.3)<br>12 (8.5)<br>7 (5.0)<br>3 (2.1)<br>3 (2.1)<br>1 (0.7)<br>1 (0.7) | Month 1<br>Month 6<br>Month 12<br>Month 24 | 40 (28.4)<br>34 (24.1)<br>42 (29.8)<br>25 (17.7) |
| 3. | New Infection (↑ in Active cysts in Location) | ↑ | 2 (0.1)<sup>†</sup> | Frontal<br>Basal Ganglia & Internal Capsule | 1 (50.0)<br>1 (50.0) | Month 1<br>Month 12 | 1 (50.0)<br>1 (50.0) |
| 4. | Image Switch from MRI to CT in Month 24 (↑ in Calcified cysts in Location) | No Change or ↑ | 14 (48.3)<sup>§</sup> | Frontal<br>Parietal<br>Occipital<br>Temporal<br>Basal Ganglia & Internal Capsule<br>Posterior Fossa | 4 (28.6)<br>4 (28.6)<br>2 (14.3)<br>1 (7.1)<br>1 (7.1)<br>1 (7.1) | Month 24 | 14 (100) |
|  |  |  |  | IV Ventricle | 1 (7.1) |  |  |
| 5. | **Patterns that cannot be easily explained: Increase in cyst of any phase with no change in image device** | No Change or ↑ | 113 (19.6)<sup>¶</sup> | Parietal | 37 (32.7) | Month 1 | 13 (11.5) |
|  |  |  |  | Temporal | 27 (23.9) | Month 6 | 36 (31.9) |
|  |  |  |  | Frontal | 18 (15.9) | Month 12 | 48 (42.5) |
|  |  |  |  | Basal Ganglia & Internal Capsule | 11 (9.7) | Month 24 | 16 (14.2) |
|  |  |  |  | Posterior Fossa | 5 (4.4) |  |  |
|  |  |  |  | Occipital | 7 (6.2) |  |  |
|  |  |  |  | Lateral Ventricles | 3 (2.7) |  |  |
|  |  |  |  | Sylvian Fissure Cistern | 2 (1.8) |  |  |
|  |  |  |  | Other Cisternal Locations | 3 (2.7) |  |  |
| 5a. | ↑ Calcified cysts with no change in image device | No Change or ↑ | 66 (58.4)<sup>#</sup> | Parietal | 22 (33.3) | Month 1 | 9 (13.6) |
|  |  |  |  | Temporal | 15 (22.7) | Month 6 | 22 (33.3) |
|  |  |  |  | Frontal | 11 (16.7) | Month 12 | 27 (40.9) |
|  |  |  |  | Basal Ganglia & Internal Capsule | 7 (10.6) | Month 24 | 8 (12.1) |
|  |  |  |  | Posterior Fossa | 3 (4.5) |  |  |
|  |  |  |  | Occipital | 3 (4.5) |  |  |
|  |  |  |  | Lateral Ventricles | 2 (3) |  |  |
|  |  |  |  | Sylvian Fissure Cistern | 2 (3) |  |  |
|  |  |  |  | Other Cisternal Locations | 1 (1.5) |  |  |
| 5b. | ↑ Transitional cysts with no change in image device | No Change or ↑ | 30 (26.5)<sup>#</sup> | Temporal | 9 (30) | Month 1 | 2 (6.7) |
|  |  |  |  | Parietal | 8 (26.7) | Month 6 | 7 (23.3) |
|  |  |  |  | Frontal | 5 (16.7) | Month 12 | 14 (46.7) |
|  |  |  |  | Basal Ganglia & Internal Capsule | 3 (10.0) | Month 24 | 7 (23.3) |
|  |  |  |  | Posterior Fossa | 2 (6.7) |  |  |
|  |  |  |  | Lateral Ventricles | 1 (3.3) |  |  |
|  |  |  |  | Other Cisternal Locations | 1 (3.3) |  |  |
|  |  |  |  | Occipital | 1 (3.3) |  |  |
| 5c. | ↑ Active cysts with no change in image device | No Change or ↑ | 17 (15.0)<sup>#</sup> | Parietal | 7 (41.2) | Month 1 | 2 (11.8) |
|  |  |  |  | Temporal | 3 (17.6) | Month 6 | 7 (41.2) |
|  |  |  |  | Occipital | 3 (17.6) | Month 12 | 7 (41.2) |
|  |  |  |  | Frontal | 2 (11.8) | Month 24 | 1 (5.9) |
|  |  |  |  | Basal Ganglia & Internal Capsule | 1 (5.9) |  |  |
|  |  |  |  | Other Cisternal Locations | 1 (5.9) |  |  |

↑ Indicates an increase, ↓ indicates a decrease
<sup>‡</sup> Denominator is the total number of scans with pattern (i.e., row percents).
<sup>†</sup> Denominator is total number of scans (n = 1372).
<sup>‡</sup> Denominator for subtypes of reverse transitions percents is the total number of reverse transition scans (n = 31).
<sup>§</sup> Denominator is number of month 24 scans where image device was switched between month 12 and month 24 (n = 49).
<sup>¶</sup> Denominator is number of scans where image device was not changed (n = 576).
<sup>#</sup> Denominator for subtypes of abnormal scans without an easy explanation is the total number of abnormal scans without an easy explanation (n = 113).

Overall, 141 scans (10.3%) demonstrated evidence of possible cyst migration (i.e., increase in the total number of cysts in the specific location being followed but no change in the total number of cysts in the brain). Most of these increases occurred in the parietal (29.8%), temporal (23.4%) and frontal lobes (16.3%), with 28.4% occurring at month one, 24.1% at month six, 29.8% at month 12, and 17.7% at month 24 of follow-up.

There were two cases in of possible new infections (new active cysts in the specific brain location being followed and an increase in the total number of active cysts in the brain). One occurred in the frontal lobe at month one and one in the basal ganglia and internal capsule region of the brain at month 12.

A total of 14 of 49 scans (48.3%) for patients that switched from MRI to CT between months 12 and 24 showed an increase in calcified cysts scan after switching. Many of these new calcified cysts appeared in the frontal (28.6%) and parietal (28.6%) lobes, with the rest evenly distributed among the occipital and temporal lobes, basal ganglia and internal capsule, posterior fossa and IV ventricle.

We observed 113 of 576 scans (19.6%) with abnormal patterns not easily explained (i.e., increases in cysts in locations with baseline total of one cyst and image device did not change between visits). These patterns were subdivided by phase where 58.4% of scans demonstrated an increase in calcified cysts, 26.5% for transitional, and 15.0% for active cysts. These patterns most often occurred in the parietal (32.7%), temporal (23.9%) and frontal lobes (15.9%) and were visible most frequently in month 12 (42.5%).

## Discussion

We found that 53.1% of the 145 patients with neurocysticercosis included in these analyses demonstrated one or more unusual patterns of cyst evolution on at least one of their four follow-up scans taken over 24 months. The most common pattern was consistent with possible cyst migration, indicated by an increase in the number of cysts in brain locations with a single cyst at baseline coupled with no change in the number of cysts in the brain, overall. This occurred in 65 (44.8%) of participants and 141 (10.3) of scans. Cyst migration has been describe before,[2,30] but our data suggests it may be more common than expected. We also found evidence suggesting reverse transitioning in 24 (16.6%) participants and 31 (2.3%) of scans, which has also been described for 14 cysts in a previous cyst-level analysis using data from this same RCT.[5] In the current study, we expanded the definition to condition on both a change in the location being followed (i.e., the total number of cysts in a later phase decreased with an increase in the number of cysts in an earlier phase) and no change in the overall brain burden. These findings also suggest that this pattern may be more common than originally thought. The number of apparent new infections in our sample was low, only observed in two (1.4%) participants in two scans (0.1%). This is consistent with a previous study that found re-infections rates to be low (0.5-3.7%) when using serology testing to identify new infections.[32] Finally, we found 12 (8.3%) patients with the appearance of new calcified cysts in 14 (48.3%) of scans related to a change from MRI to CT scan. This suggests that a change in imaging type likely results in a change in the patient’s NCC cyst profile in radiology reports. We also found unexpected increases in the number of calcified, transitional, and active cysts unrelated to a change in imaging device. These seem likely to be related to factors we were unable to assess such as new cysts becoming visible as swelling in the brain and edema subsided. Of course, it is possible that all or a large proportion of these unexpected cyst evolution patterns are due to errors in the radiologist reading of the scans, which, as we noted before, is known to have fairly poor reliability.[4,23–25]

In our study, cyst evolutionary patterns were identified for the most part by following cysts in locations where the cyst burden at baseline was one. While this was useful in trying to understand cyst evolution, these individual cysts could not be followed over time as, in many cases, the number of cysts in that location increased to more than one over time. This could be due to new infections, cyst migration, change in the form of imaging, or the appearance of new cysts for other reasons not easily explained. Even if the number of cysts in a given location did not increase beyond one, the assumption that we are following the same cyst could be incorrect. For example, consider reverse transition patterns where it is conceivable that a cyst did not reverse transition, but instead the original cyst resolved and a new cyst in an earlier phase appeared due to a new infection, cyst migration, change in imaging, etc. Thus, while the patterns we describe may be consistent with potential and understandable explanations, there are alternate explanations, which we cannot ignore and present in Table 1 for consideration. Without a cyst-level reading of scans that attempts to follow individual cysts over time, it is not possible to rule out multiple explanations for a given pattern. Even with a radiologist actively following individual cysts, it may still be impossible to document and understand the evolution of each cyst. These are the challenges in understanding NCC cyst evolution, even when attempting to follow individual cysts over time. Given that most data on NCC cysts is collected and followed in aggregate,[4,11,13,17,18] the current paradigm of NCC research makes it unlikely that we will understand both how cysts evolve and the true reasons behind the patterns we describe in this paper.

This study is the first to describe unexpected NCC cyst patterns over time and explore potential explanations for these patterns. Future research on NCC patients should include cyst-level readings of the scans to improve our understanding about NCC evolution (i.e., can cysts truly reverse transition), cyst migration, if new infections or gemmation occurs, and if new cysts become apparent on imaging when edema is reduced and/or the form of imaging changes. With an improved understanding of NCC cyst evolution, we can better evaluate disease progress in relation to treatment and time – which is key to the development of new, more effective treatment. The current study has several limitations that should be considered when interpreting the findings. Some patient follow-up scans were missing (n = 518, 27.4% of total possible scans); in some cases, the time interval between scans was longer than planned. In addition, there may have been measurement error since radiology readings are not highly reliable. In some cases, our underlying assumptions about what each pattern likely indicated could be incorrect. Finally, the data come from a study conducted among patients in Ecuador in 2001-2005. It could be that the distribution of cyst patterns we describe here is different in other populations and may have changed over time such that a similar study using more recent data would have different results. Thus, the findings we present here should not be generalized and more recent NCC research is needed in Ecuador and other populations. Despite these limitations, this is the first study to our knowledge to systematically describe unusual patterns in NCC cyst evolution over time. These findings may be useful to inform how radiologists read images of NCC patients and may generate ideas for future research to help us better understand this infectious disease.

## Data Availability

Data not available.

## Author Bio

Meghana G. Shamsunder is a doctoral candidate at CUNY’s School of Public Health and Health Policy in the Department of Epidemiology and Biostatistics. Her research focuses on advanced methods in epidemiology.

